# Autonomy-Supportive Sexual Health Communication and Sexual Health Behaviors for Black and Latino MSM in the House Ball Community: Protocol for a Social Network Analysis and Exploratory Structural Equation Model

**DOI:** 10.1101/2022.10.06.22280768

**Authors:** Martez D. R. Smith, Natalie M. Leblanc, LaRon E. Nelson, James M. McMahon

## Abstract

**Background:** Black and Latino men who have sex with men (MSM) have the highest risk of HIV of any group in the U.S. Prevalence could be even higher among Black and Latino MSM belonging to the House Ball Community (HBC), an understudied community comprised primarily of Black and Latino sexual and gender minorities, with HIV prevalence from non-probability samples ranging from 4% to 38%.

**Objectives:** Basic Psychological Needs Theory will be utilized to understand how sexual health communication (SHC) influences sexual health behaviors of HBC-MSM. The proposed study will advance this goal by describing characteristics of SHC embedded within social and sexual networks, and identifying the effects of SHC on sexual health behaviors among a sample of Black and Latino HBC-MSM.

**Methods:** This study entails cross-sectional quantitative survey design with internet-based data collection to test a theory-driven model of the effects of autonomy supportive communication on sexual health behaviors. Respondent-driven sampling (RDS) and internet driven sampling (ID) will be used to recruit a sample of 200 HBC-MSM. We will utilize egocentric network analysis to describe (a) the HBC-specific social and/or sexual network members who provide SHC; (b) the degree of autonomy support provided by network members, and (c) the sexual health behaviors characterizing the sample of HBC-MSM. Structural equation modeling (SEM) will be performed to test associations between autonomy supportive sexual health communication (independent variable) and sexual health behaviors (outcome), with needs satisfaction and intrinsic motivation as intervening mediators.

**Discussion:** Such knowledge is necessary to expand understanding of how SHC impacts sexual health behaviors for HBC-MSM. The study provides an critical perspective on sexual health behaviors and motivations as participants operate in HBC. Knowledge generated from this study will help improve current HIV prevention interventions, as well as inform the development of future interventions, tailored to HBC-MSM.

**Trial registration:** N/A

## BACKGROUND

Black men who have sex with men (MSM) are increasingly disproportionately impacted by HIV, accounting for 62.7% of all new HIV diagnoses among Black Americans and 26% of all diagnoses in the U.S. in 2020 (1). Similarly, Latino MSM account for 77% of all new HIV diagnoses among Latino Americans and 20% of all diagnoses in the U.S (1). Projections indicate that 1 in 2 Black MSM and 1 in 4 Latino MSM will be diagnosed with HIV during their lifetime if current trends persist (2). Understanding the social and structural context of HIV risk among Black and Latino MSM is crucial to developing and implementing interventions that effectively address the HIV epidemic among this population. To more fully understand the social and structural contexts of HIV risk among Black and Latino MSM, we must examine an understudied, yet crucial aspect of their social life – participation in a form of chosen kinship bonds known as the House Ball Community (HBC). Empirical estimates are lacking as to the proportion of MSM who will become involved in the HBC at some stage of their lives; however, in a 2017 study of MSM in New Orleans, Zarwell and Robinson found that 173 of 553 participants (31%) reported belonging to a house, gay family, or pageant house (3). An important concern is that HBC-MSM may be disproportionately impacted by new HIV diagnoses in comparison to Black and Latino MSM broadly (4-8).

National estimates of HIV prevalence among Black and Latino MSM range from 18% to 28% (9, 10). HIV prevalence among HBC-specific largely Black MSM samples range from 4% to 38% (4-8). In a study of 361 HBC members (including 301 MSM) in New York City (NYC) who underwent HIV testing, 21% received a positive result of whom 75% were newly diagnosed (5). The majority of the sample (53%) were Black MSM, while Latino MSM accounted for 44% of the sample. Black MSM had an HIV infection rate 8 times higher than the other groups in the sample (5, 11). Another Chicago-based study among MSM yielded a higher HIV prevalence among HBC-MSM when compared to non-HBC members (38% vs. 33%)(7). This evidence demonstrates a clear disparity in HIV infection rates within sub-communities of MSM. Although no theories have been forthcoming to elucidate the disproportionate HIV infection rates of HBC-MSM when compared to MSM broadly, the age demographic of HBC-MSM may explain some of the disparity in HIV diagnoses. The mean age for HBC members tends to be young, ranging from 20 to 25 years (4-8, 11). Black MSM, ages 13-34, and particularly Black and Latino MSM ages 25-34, have much higher rates of HIV infection (1), particularly because HIV prevalence is higher within Black, Latino, gay and bisexual communities (1, 12) and respective sexual networks tend to be racially homogenous (13-15). Additionally, a suite of interrelated biological, individual behavioral, and structural and institutional factors coalesce to exacerbate Black and Latino MSM odds of contracting HIV (16).

The HBC is a chosen kinship network (17, 18) that consists of different “houses” whose primary purpose is to groom and mentor house members for participation in elaborate competitions against other houses, known as balls. Houses compete in balls across the U.S. and globally (18-21). It is important to note that houses are not physical locations for their members, rather, family-like social structures that serve as a social home for community members (11, 21). Successful competition in balls affords a particular house and the individual house member notoriety, elevated social status in the HBC at-large, trophies, and cash prizes. Houses, similar to other forms of chosen kinship, are constructed of mutually chosen kinship ties that mimic relationships and roles found in the traditional western nuclear family (21). For example, house managers are referred to as parents (e.g. mothers and fathers, based upon gender identity/expression) who oversee house members (known as “children”). Grooming and mentorship for participation in ball competitions includes hosting practices for house members, advising members on what categories to compete in, and advising house members on how to compete successfully (21, 22). The grooming and mentorship provided by house parents often expands beyond the grooming for ball competitions into other aspects of house children’s lives - such as personal and professional aspects. Membership in a house often affords Black MSM a sense of camaraderie, social support, and a platform through which artistic expression and productive competition can occur (4-8). As with other kinship ties and family relationships, the social bonds established via HBC membership provide a rich social context through which health messaging, including sexual health messaging, is communicated.

HBC-MSM face a complex dynamic in relation to HIV transmission and vulnerability to infection. Yet the social support and comradery afforded to HBC-MSM via fellow house members and ball participants has the potential to mitigate the disproportionate number of HBC-MSM impacted by HIV. Various forms of social support (e.g. encouraging safe sex and/or HIV testing, providing money or a place to stay, and/or providing emotional support) from HBC peer network members have been positively correlated with sexual health behaviors (4, 6, 23). Among participants who received higher levels of social support for HIV testing and safe sex, decreased odds of CAI and reduced instances of delayed HIV testing were documented (6). Despite the potential sexual health promotion that HBC-MSM may benefit from, threats to sexual health persist such as CAI (6, 23), negative social influences on sexual behaviors from non-HBC and non-house members (4, 23), and increased sexual risk behavior when social networks overlap (4, 6, 23). Basic Psychological Needs Theory (BPNT), a sub-theory of Self-Determination Theory (24), provides a useful framework for testing a hypothesis to explain these findings; autonomy-supportive communication within the HBC invokes greater motivation for heath behavior by supporting autonomy.

BPNT posits that fulfilment of three basic psychological needs (autonomy, relatedness, and competence), increases intrinsic motivation and promotes optimal development, integration, and wellbeing throughout the life course (25). *Autonomy* can be understood as volition, or an individual endorsing a behavior, task, or activity at their free will. *Relatedness* is understood as the social connection between individuals within the context of a specific behavior, task or activity. *Competence* is understood as information or knowledge of a certain subject matter. The fulfilment of autonomy needs facilitate fulfilment of the other two needs (26, 27). The fulfilment of these basic psychological needs (needs satisfaction) facilitates intrinsic motivation to engage in health-promoting activities, as well as achieve favorable psychological performance as a result of engaging in those activities. According to BPNT, significant/influential peers and authoritative figures who provide support for and facilitate fulfilment of autonomy needs will facilitate fulfilment of relatedness and competence needs (25). Research has shown that autonomy support and fulfillment of basic psychological needs are associated with positive sexual health behavior and HIV-related outcomes in both adolescent and adult populations (28-36). Thus, this study will apply BPNT to investigate autonomy supportive sexual health communication with HBC-MSM that may have a direct impact on sexual health behaviors through needs satisfaction and intrinsic motivation. These theory-derived mechanisms have not been previously studied within the context of chosen kinship structures fostered in HBC.

### Study Rationale and Aims

The proposed study will investigate three specific aims:

#### Aim 1

Describe (a) the social and sexual networks of HBC-MSM; (b) assess the degree of autonomy supportive sexual health communication provided by social and sexual network members, and (c) describe the sexual health behaviors characterizing the sample of HBC-MSM.

#### Aim 2

Assess a theoretical model in which (a) autonomy supportive sexual health communication has a direct effect on sexual health behaviors; (b) intrinsic motivation mediates the relationship between autonomy support and sexual health behavior; and (c) needs satisfaction mediates the relationship between autonomy support and intrinsic motivation.

**H2a**: Greater autonomy support for sexual health will be positively associated with sexual health behaviors <u>(recent HIV/STI testing, fewer sexual partners, consistent condom use, engagement in care, and PrEP/ART adherence).</u>
**H2b**: Autonomy support will be positively associated with greater intrinsic motivation, which, in turn will be associated with greater sexual health behavior.
**H2c**: Autonomy support will be positively associated with increased needs satisfaction, which, in turn will be associated with greater intrinsic motivation for sexual health behaviors.

## METHODS

### Study Design

This study will entail a cross-sectional quantitative survey design to test a theory-driven model of the effects of autonomy supportive communication on sexual health behaviors among Black and Latino HBC-MSM. Egocentric network analysis will be utilized to describe (a) the HBC-specific social and/or sexual network members who provide sexual health support; (b) the degree of autonomy support provided by network members, and (c) the sexual health behaviors characterizing a sample of HBC-MSM. Structural equation modeling (SEM) will be performed to test associations between autonomy supportive sexual health communication (independent variable) and sexual health behavior (outcome), with needs satisfaction and intrinsic motivation as intervening mediators (see Fig. 1).

**Fig 1.**
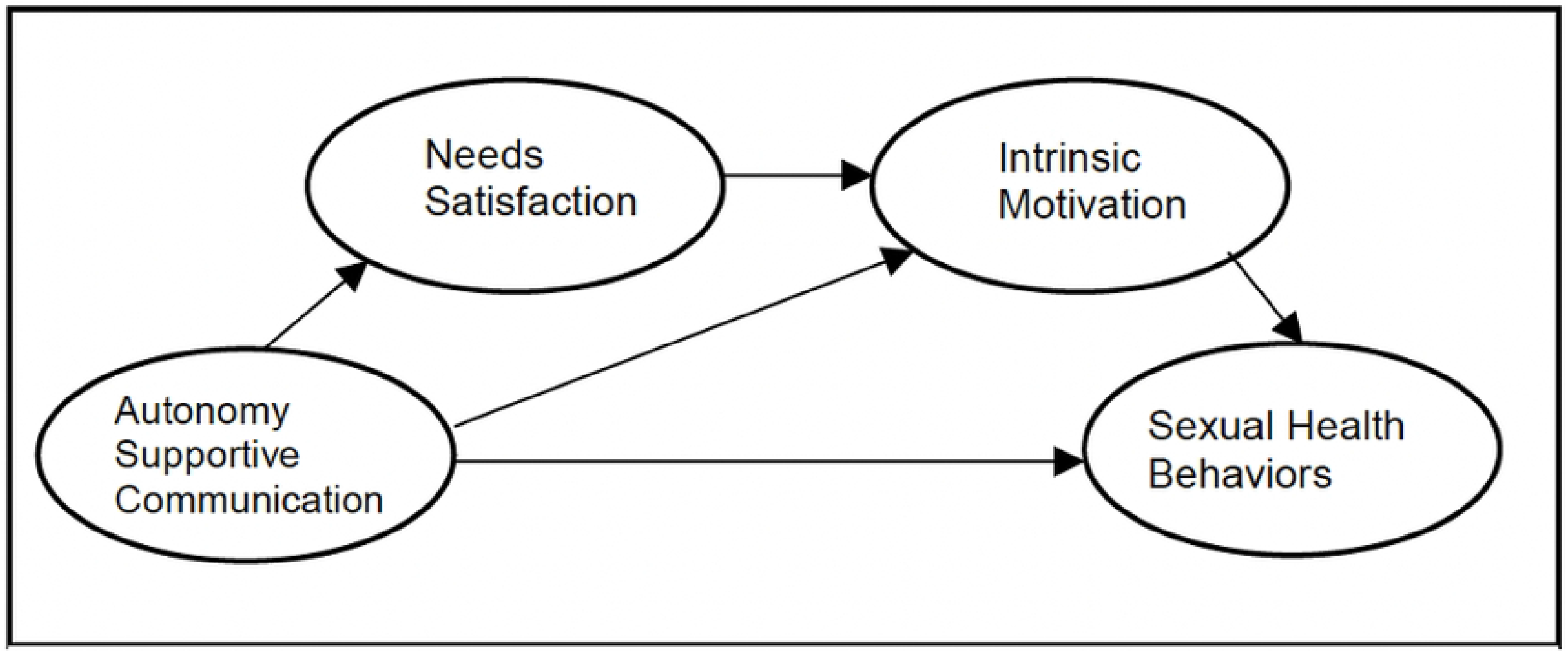
Hypothesized SEM for Direct and Indirect Effects of Predictor Variables on Sexual Health behaviors.

### Study Setting

Recruitment for the study will be conducted in the New York City (NYC) metropolitan area, which is the geographic area with the highest concentration of individuals in the HBC. Participants who reside outside of the NYC metro area are eligible so long as they reside in the United States. Participants who are eligible and choose to enroll in the study will have the option of completing the survey at their own leisure from their personal electronic devices that have internet access and web browsing capabilities.

### Study Participants

#### Eligibility

Participants must meet the following eligibility criteria:

1. 18 years of age or older;
2. self-identified Black, African American, Latino or Hispanic race or ethnicity;
3. fluent in English;
4. a working personal email address;
5. assigned male at birth;
6. currently identify as a man;
7. report sexual contact (oral, anal or otherwise) with a male in the previous year;
8. self-reported member of the HBC; and
9. reside within the United States

#### Ethics Approval

This study was approved by the University of Rochester Research Subjects Review Board (RSRB) (United States) on June 17, 2021: STUDY00005823

### Study Measures

#### Aim 1

To capture individual-level data on social and sexual network members each participant will be asked to list the individuals in their social and sexual networks who they have had sexual health-related discussions with in the previous 30 days. Respondents will be asked to characterize each of their identified network members on a series of attributes, including whether they are sexual partners and/or fellow HBC house members, and if so, their role in the house, degree of personal influence, age, race/ethnicity, and to what degree the sexual health-related discussion was perceived as supportive of participants’ autonomy (37).

#### Aim 2

Dependent variables: Five separate SEM models will be assessed corresponding to 5 status-neutral sexual health behavior outcomes: (1) STI and HIV testing, (2) number of sexual partners, (3) condom use, (4) engagement in care and (5) antiretroviral adherence (ART or PrEP).

**DV1)** Recent HIV/STI testing will be an observed dichotomous variable, operationalized as a test that occurred within the past six months.

**DV2)** Number sexual partners will be a count variable captured by a single-item self-reported count of partners during the last 30 days. Sexual partners are defined as any individual whom participants have had sexual contact with (oral, anal or otherwise).

**DV3)** A composite score of condom use consistency for romantic partners will be taken using a 4-item survey; participants will be asked about condom used during last sex with primary and non-primary sexual partners (“Did you or your partner use a condom the last time you had sexual intercourse?”) and participants will be asked about condom use consistency with non-romantic sexual partners in the past 30 days using a 7-point Likert-type scale with answers ranging from “I always use condoms with my partner(s)” to “Never” (38). Additional items will inquire about sex type (e.g. insertive/receptive oral and receptive/insertive anal).

**DV4)** Engagement in HIV-specific care will be measured as a dichotomous variable, indicated by any consultation about PrEP with a provider (for participants who are HIV seronegative), or having attended a doctor’s appointment within the last 6 months (for participants who are HIV seropositive).

**DV5)** For people who are HIV-negative, PrEP adherence will be captured by a single-item measure of percentage of medication taken during the past 4 weeks, “Thinking about the past 4 weeks, how often did you take all of your PrEP medications?” (39). For people living with HIV, adherence will be captured by an observed continuous single-item, “What percent of the time were you able to take your medications exactly as your doctor prescribed them?” Response categories for PrEP and ART adherence will range from 0% to 100% in increments of 10% (40, 41)(Fig. 1).

### Independent variable

Autonomy supportive sexual health communication will be measured as a latent variable using an adapted version of the 21-item Perceived Autonomy Support Scale for Romantic Partners (PASS-RP). Only 9 items that load onto the autonomy support factor will be used. The PASS-RP will be modified to measure autonomy support in aggregate provided by HBC-MSM social network members. Prior research has demonstrated high internal consistency (α>0.89) and convergent and divergent validity confirmed via correlational patterns with the Psychological Control Scale, as well as monitoring and acceptance domains of parenting (42).

### Needs Satisfaction

Needs satisfaction will be measured utilizing The Basic Psychological Need Satisfaction Scale in General (BNSG-S), a 21-item Likert-scale tool (25, 43). The BNSG-S is comprised of three subscales which measure each of the BPNs (autonomy, relatedness, and competence). Scores for each subscale indicate the degree to which the BPN is satisfied in the individual. The autonomy subscale contains 7 items, the competence subscale contains 6 items, and the relatedness subscale contains 8 items. A study on the empirical dimensionality of the BNSG-S found subscale reliability values were 0.60 for autonomy, 0.55 for competence and 0.78 for relatedness. The subscales were externally validated by testing the hypothesis that increased subscale scores would be positively associated with greater scores on various domains of wellbeing (43).

### Intrinsic Motivation

A modified version of the Treatment Self-Regulation Questionnaire (TRSQ), a 15-item, seven-point Likert-type scale, will be used to measure intrinsic motivation for sexual health promotion and behavior. The measure consists of 4 subscales, each reflective of one of 4 types of motivation. The amotivation subscale contains 3 items, the external regulation subscale contains 4 items, the introjection subscale contains 2 items, and the autonomous regulation subscale contains 6 items. The TRSQ contains one stem statement to which participants respond. For this study, the stem will be modified to assess motivation for protecting one’s self from sexually transmitted infections, including HIV. For example, the statement, “the reason why I would try to protect myself from getting HIV/STIs is:” followed by the 15 items. TRSQ has been validated in numerous contexts for various health behaviors (e.g. diet, exercise, and tobacco use) with an internal consistency value α>0.73 for most subscales. Analyses of invariance across three health behaviors captured at three different sites provide evidence for validity of the TRSQ. Higher overall scores indicate a greater intrinsic motivation (44).

### Covariates

Several variables that have been found to influence sexual health behaviors for Black MSM will be entered into the model, including: age (45, 46), race (45), stimulant use immediately before or during sex (46-48), income (46), years of education (47), employment status (49), homophobia (50, 51), HIV-related stigma (51), relationship status (52), housing instability (46, 52).

### Recruitment

A novel, hybrid sampling approach of previously utilized and validated methods to recruit hard-to-reach populations will be employed integrating respondent-driven sampling [RDS], time-location sampling [TLS)] and internet-driven sampling [ID] (53, 54). A small, semi-probabilistic sample of subjects, known as “seeds” (wave 1), will be recruited via TLS and ID methods. Spaces (location) as well as days and times (time) of target population activity will be identified to construct a sampling frame of time-location (TL) units. TL units will be selected at random and visited by the researcher, who will disseminate study flyers. A subset of TLS venues will consist of HBC-sponsored events. The study will be advertised via fliers circulated during in-person and virtual HBC events, including virtual balls. The flier will contain a quick response (QR) code and link that will direct potential subjects to a landing webpage, which will include a brief description of the study. Potential subjects will have the option to continue to the screening survey by clicking “submit” at the bottom of the landing webpage. Subjects will not be compensated for taking the screener survey. ID methods will consist of posting study ads in digital spaces where HBC members convene and exchange information (e.g. Facebook groups). An adaptive approach will be utilized so that additional recruitment techniques will be implemented to ensure that our targeted N=200 is reached.

Once each subject completes the study, they will have the option to refer up to three of their HBC-social network members in the study. Subsequently the nominated social network member will receive an automated email from the PIs. The email will contain instructions on how to access the study webpage and screener. Evidence suggests that recruiting 4 to 6 waves of subjects will ensure accurate parameter estimates for hard-to-reach populations (55, 56). For this study, enrollment of 25 “seed” subjects with an average referral of 1.5 peers per subject will yield a total sample size of 203 over 4 waves of referrals.

### Enrollment and Data Collection

After completing the eligibility screener, subjects will then be asked if they would like to enroll in the study. If they select yes, they will proceed to the information sheet. Upon review and consent with the information sheet, participants will proceed to the review instructions for completing the survey. Survey measures consist of items inquiring about subjects’ history, including sexual behavior, substance use, sexual health status (HIV and STI diagnoses), and conversations around sexual health. Upon completion of the survey, a completion page will be displayed thanking the participant for their time. Additionally, participants will be required to provide their first and last name, email address, phone number, and complete a CAPTCHA (Completely Automated Public Turing to tell computers and humans Apart) to verify identity and facilitate compensation. Each record will be reviewed for validity of responses prior to distributing compensation (see the Data Validation protocol in S1). Compensation will be sent electronically via email after completing data collection and validation processes.

Participants will have the option of providing up to 3 email addresses for peer referrals. Each peer-referred participant survey will be automatically linked in REDCap to the seed referral survey via referral ID. Referred individuals will receive an automated email containing a customized link to the study webpage and screener.

### Power and Sample Size

We will enroll 200 participants in this study. Based on α =.05, power of 80%, and an effect size of Cohen’s d=.30, the minimum sample size needed to detect an effect for a given SEM path is N=200 (57-59). This sample size is based on a final hypothesized model consisting of 3 latent variables (autonomy supportive sexual health behavior communication, needs satisfaction, intrinsic motivation) and observed sexual health behavior outcomes.

### Statistical Analysis Plan

Appropriate data cleaning methods will be applied prior to descriptive and inferential analysis, including

missing data handling, multicollinearity, and violations of model assumptions.

#### Aim 1

Egocentric network analysis will be used to describe (a) the HBC-specific social and/or sexual

network members who provide sexual health support; (b) the degree of autonomy support provided by network

members, and (c) the sexual health behaviors characterizing a sample of HBC-MSM. Descriptive statistics

will be utilized to characterize attributes of participant networks, including sexual health communication

patterns and the amount of autonomy supportive and sexual health behaviors embedded within participant

social networks. Visualizations of network data will be generated via EgoNet to identify patterns in network

structure, and to obtain general information about social networks including ego-alter ties (network size and

multiplexity), in addition to alter attributes (composition, ego-alter similarity, heterogeneity/diversity) (37). Network size refers to number of individuals within a network. Multiplexity indicates the number of roles or ties that define a particular relationship. For example, a multiplex tie between an ego and alter would be when the tie is representative of being a co-worker and a friend with someone (multiple roles/connections). Ego-alter similarity consists of 3 mechanisms: 1) preference; individuals generally socialize and bond with individuals who are similar to them (homophily, measured by demographic characteristics including race, ethnicity, age, and HBC participation/membership) 2) availability of social connections; people generally socialize and bond with individuals they come into physical contact with 3) Influence (measured by a single-item, Likert scale); individuals become increasingly homophilous over time via repeated social interactions. Influence can be utilized to conceptualize and measure various social network characteristics such as the benefits of homophily (identify affirmation, insulation from outside influences) as well as measuring influence over time. Heterogeneity/diversity is measure of how demographically similar altars are to each other instead of egos (37).

#### Aim 2

Structural equation modeling (SEM) will be performed using RStudio and the Lavaan add-on package for latent variable modeling to assess a theoretical model (see Fig. 1). The SEM will assess the following model-derived hypotheses: autonomy supportive sexual health communication will have a direct positive effect on HIV testing, engagement in care, and ARV adherence and a direct negative effect on CAI and number of sexual partners; Autonomy supportive sexual health communication will have a negative indirect effect on CAI and number of sexual partners through intrinsic motivation.

#### Measurement model

The measurement component of the SEM will first be evaluated using exploratory factor

analysis (EFA) to examine the underlying correlational structure of each latent variable, in order to verify

modifications involving individual items and theoretical subscales (60, 61). Confirmatory factor analysis (CFA)

will then be performed to assess the overall measurement model based on path coefficients and standard

model fit indices. CFA results may lead to further modification of the measurement model (62).

#### Structural model specification

Missing data will be addressed for all models via full information maximum likelihood (FIML) estimation (63). SEM models will be performed using weighted least squares with missing values estimator and mixed linear, logistic, and probit link functions (64). Structural models involving pairs of latent variables will first be performed to assess the bivariate relationships among variables.

#### Model identification

Model complexity will be considered when specifying the multivariable structural models in order to achieve model identification (62). Thus, the full model will be specified in stages of increasing model complexity.

#### Model estimation

The direct effects of autonomy supportive sexual health communication on sexual health

behaviors will be assessed. Next, the latent needs satisfaction variables will be added as mediators between

autonomy supportive sexual health communication and the selected latent intrinsic motivation variables.

Subsequently, the relationships between the specified intrinsic motivation latent variables and sexual health

behaviors will be assessed.

#### Full model specification

Finally, the full models, each based on 1 of 5 outcome variables will be assessed. Based on our conceptual model, we expect to observe path coefficients demonstrating conclusive effects (i.e., 95% confidence intervals not containing the null value). Overall model fit will also be assessed using the following model fit indices: Absolute fit indices, model chi-square, root mean square error of approximation (RMSEA), goodness-of-fit (GFI), adjusted goodness-of-fit (AGFI), root mean square residual (RMR), standardized RMR statistics. Traditional cutoff criteria will be used for fit indices: non-significant chi-square test, RMSEA values less than 0.08, and GFI/AGFI less than or equal to 0.05 (64).

### Ethical Considerations

#### Process of Consent/Assent

Since this study is an online-based, one-time survey, an information sheet will be used instead of requiring signatures on an informed consent document. Screening, enrollment and data collection will take place online and data will be stored in REDcap. Coercion will be minimized by emphasizing that subjects can withdraw from the online study procedures, including screening, consent, or survey measures at any time. This text will be emphasized in the information sheet, which will also include a button that allows subjects to save and print the form for their records.

#### Confidentiality & Security of Data

To maintain primary and confidentiality of subject and research data we will be foregoing the paper data collection process (eliminating opportunities for lost or stolen items with identifying information), and storing all identifying information on a password protected encrypted server that only the research team will have access to. All data will be de-identified during the data cleaning process that will take place after data collection ceases.

To protect against potential confidentiality breaches via internet, online data collection will be facilitated utilizing REDCaps, a University of Rochester Research Subjects Review Board-approved survey software. All datasets will be password protected. Identifiers will be removed from each subject survey and will be replaced with a unique study identifier code prior to data analysis.

#### Data Storage and Access

Data will be stored and managed via an academic REDCap system. Coded data will be stored separately from identifiable data on a secured server and will only be accessible via in-person connection on campus or via encrypted, firewall-secured, virtual private network. Outputs from analyses will be password protected and stored in an electronic folder separately from the coded and uncoded datasets. The study team will have access to the coded data and outputs. All data will be deleted 3 years after the completion of the study.

#### Data and Safety Monitoring Plan

The study team will meet to conduct data monitoring on a monthly basis to ensure integrity. No stopping rules apply since this is not a clinical trial. All demographic data and survey data will be reviewed to ensure completeness and safety. No adverse events or stopping rules apply since this is not a clinical trial. If fraud, or some other type of adverse event is suspected, the investigative team will work together to submit a written notice of such activity to RSRB for further guidance.

#### Reporting of Adverse Events

This study protocol presents minimal risk to participants and adverse events are not anticipated. In the occurrence of an adverse event or an *unanticipated problem involving risks to subjects or others*, written notice of the event should be submitted to the RSRB within 48 hours of being identified by the study team. The University of Rochester Office for Human Subject Protection (OHSP) will provide further guidance upon notification, as indicated in OSHP Policy 801 - *Reporting Research Events*.

#### Management of Interim Results

Authors MDRS and JMM will be responsible for the implementation of the Research Strategy and specific aims and will ensure that mechanisms are set in place to ensure institutional compliance with US laws, DHHS and NIH policies including human subjects’ research, data and facilities.

#### Dissemination Plan

Abstracts for presentations on the final egocentric network analysis, as well as developed and validated models of autonomy supportive sexual health communication will submitted for scientific conferences. Manuscripts based on findings will be published in scientific, peer-reviewed journals. Results will be utilized to inform the development of a culturally-tailored sexual health behavior promotion intervention for HBC-MSM.

#### Protocol modifications

Protocol modifications will be initiated by Mr. Smith in consultation with Dr. McMahon. If additional guidance is necessary the full sponsoring team for the F31 funding the study will be convened (Smith, McMahon, Nelson, Leblanc) to discuss challenges and brainstorm solutions. The study team will consult with the RSRB and OHSP for recommendations as well.

#### Payments for Participation

Participants will be compensated with a $35 Amazon gift card that will be distributed via email after completing the study procedures when the study record is validated. Participants receive an additional $5 Amazon gift card for each eligible peer referral that completes the survey, for a total of up to $50 in Amazon gift cards.

## DISCUSSION

As a novel, vibrant and impactful community embedded in larger spheres of Black, Latino and other racially diverse LGBT communities, the HBC provides its members with social networks through which knowledge transfer and resource sharing easily occur. This study investigates the social and sexual networks of HBC-MSM and explores how autonomy supportive sexual health communication influences motivation for sexual health behaviors. For aim 1 we will collect information on social and sexual network members with whom HBC-MSM have had sexual health-related conversations within the previous 30 days. We will utilize egocentric network data to obtain general information about social networks including ego-alter ties (network size and multiplexity), in addition to alter attributes (composition, ego-alter similarity, heterogeneity/diversity). To achieve aim 2, we will test hypothesized structural equation models to identify direct and indirect effects of autonomy supportive sexual health communication on observed sexual health behaviors (Recent HIV/STI testing, number of sexual partners, condom use, engagement in care, and PrEP and ART adherence) via basic psychological needs satisfaction and intrinsic motivation for sexual health behaviors. Upon conclusion, findings will be utilized to conduct a pilot study of a peer-led, sexual health intervention for HBC-MSM.

Findings from this study will contribute to the development of a multicomponent, status-neutral HIV prevention intervention to promote 1) autonomy supportive sexual health communication and 2) fulfilment of basic psychological needs among HBC-MSM. Fulfillment of basic psychological needs facilitates intrinsic motivation for sexual health behaviors, including increased engagement in HIV prevention, treatment, and care.

### Trial status

The paper is adapted from Protocol version 1.4, approved by: University of Rochester Research Subjects Review Board (United States) on September 29, 2022.

## Data Availability

No datasets were generated or analyzed during the current study. All relevant data from this study will be made available upon study completion.

## List of abbreviations

AGFI: Adjusted Goodness-of-Fit
AIDS: Acquired immunodeficiency syndrome
ARV: Antiretroviral
BNSG-S: Basic Psychological Need Satisfaction Scale in General
BPNT: Basic Psychological Needs Theory
CAI: Condomless Anal Intercourse
CAPTCHA: Completely Automated Public Turing to tell Computers and Humans Apart
CFA: Confirmatory Factor Analysis
DHHS: Department of Health and Human Services
EFA: Exploratory Factor Analysis
FIML: Full Information Maximum Likelihood
GFI: Goodness-of-Fit
HBC: House Ball Community
HIV: Human immunodeficiency virus
ID: Internet-driven
IP: Internet Protocol
LGBT: Lesbian, Gay, Bisexual, and Transgender
MSM: Men Who have Sex with Men
NIH: National Institutes of Health
NYC: New York City
OHSP: Office for Human Subject Protection
PASS-RP: Perceived Autonomy Support Scale for Romantic Partners
PI: Principal Investigator
PrEP: Pre-Exposure Prophylaxis
QR: Quick Response
RDS: Respondent-driven sampling
REDCap: Research Electronic Data Capture
RMR: Root Mean Square Residual
RMSEA: Root Mean Square Error of Approximation
RSRB: Research Subjects Review Board
SDT: Self-Determination Theory
SEM: Structural Equation Model
STI: Sexually Transmitted Infection
TRSQ: Treatment Self-Regulation Questionnaire
UR-SON: University of Rochester School of Nursing
US: United States

## Declarations

### Consent for publication

Not applicable. No individual-level data is included in this report.

### Availability of data and materials

The datasets generated and analyzed during the current study will be de-identified and made available from the corresponding author on reasonable request.

### Trial Sponsor Contact Information

Susannah Allison, PhD, BA, Training Director and Program Officer, Division of AIDS Research, 5601 Fishers Lane, Room 9E26, Rockville, MD 20852

### Authors’ contributions

MDRS and JMM contributed to study conceptualization and design. JMM, LEN, NL, and MDRS contributed to the refinement of the study protocol. All authors read and approved the final manuscript.

## Acknowledgements

No acknowledgements.

## Authors’ information

No additional information

## Footnotes

None

